# Accounting for incomplete testing in the estimation of epidemic parameters

**DOI:** 10.1101/2020.04.08.20058313

**Authors:** R.A. Betensky, Y. Feng

## Abstract

As the COVID-19 pandemic spreads across the world, it is important to understand its features and responses to public health interventions in real-time. The field of infectious diseases epidemiology has highly advanced modeling strategies that yield relevant estimates. These include the doubling time of the epidemic and various other representations of the numbers of cases identified over time. Crude estimates of these quantities suffer from dependence on the underlying testing strategies within communities. We clarify the functional relationship between testing and the epidemic parameters, and thereby derive sensitivity analyses that explore the range of possible truths under various testing dynamics. We derive the required adjustment to the estimates of interest for New York City. We demonstrate that crude estimates that assume stable testing or complete testing can be biased.

## Introduction

The features of an epidemic are summarized and visualized using several measures. One such measure is the epidemic curve (1), which depicts numbers of reported cases as a function of time. Another is the cumulative version of this curve, which depicts total reported cases as a function of time. While these are important and of high interest for the purpose of understanding the dynamics of the epidemic and the utility of public health interventions, they are limited by the testing processes. That is, the numbers of cases identified are limited by the numbers of tests conducted. A decrease or flattening of the epidemic curve could be due to a decrease in the rate of infection, or it might be due to a decrease in testing, or some combination of the two. An alternative measure of the epidemic is its current *doubling time* (2), i.e., the time needed for the cumulative incidence to double. We suggest that the doubling time should be used as a primary measure of the epidemic due to its clear interpretation in light of testing policies and dynamics and the potential to conduct meaningful sensitivity analyses of it. Likewise, cumulative incidence curves that are normalized to recent dates share the same desirable features.

## Methods

If the epidemic follows an exponential growth model, the doubling time is constant. That is, the time for the number of cases to double remains the same at all times during the course of the epidemic. An increase in the doubling time is an indicator that the growth of the epidemic is slowing, which in turn, indicates that public health policies, such as social distancing, are displaying efficacy. For this reason, the doubling time is a useful descriptor of the epidemic.

It is important to understand how changes in testing policies and implementation might affect the estimates of doubling time, given that as for the epidemic curve, it is intertwined with testing. The true doubling time at time *t* is defined as

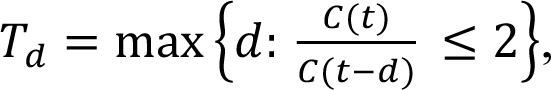

where *C(t)* is the actual total number of cases at time *t*. Because we cannot know C(t), instead we estimate the doubling time at time t as

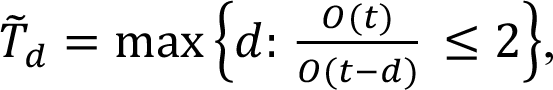

where O(t) is the observed total number of confirmed cases at time t. It is necessarily the case that O(t) ≤C(t). There is a simple relationship between O(t) and C(t), which follows from an application of Bayes theorem:

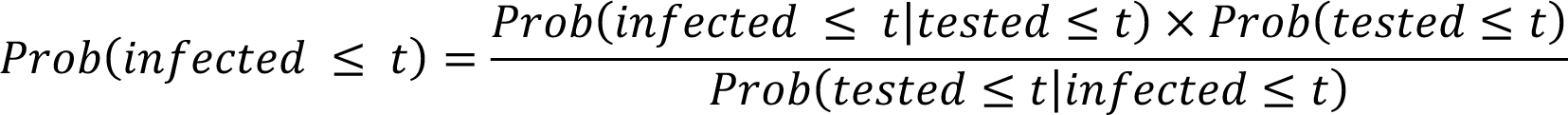

Or equivalently,

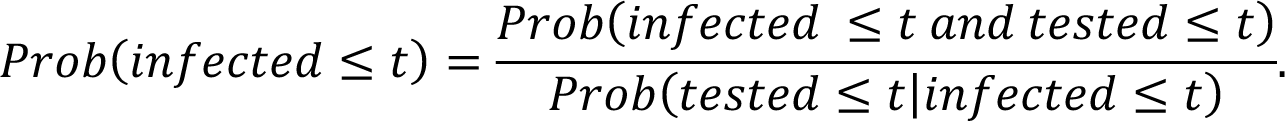

This implies that we can estimate the true number infected, C(t), using its conditional expected value given the observed number infected, E[C(t)|O(t)], which is given by:

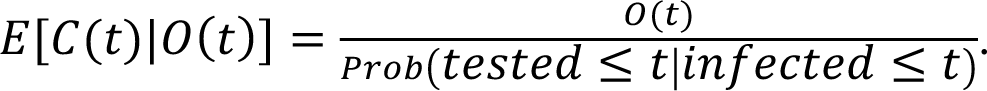

This, in turn, means that we can re-express the true doubling time as a function of the observed numbers of cases, along with the probabilities of testing of those who are infected:

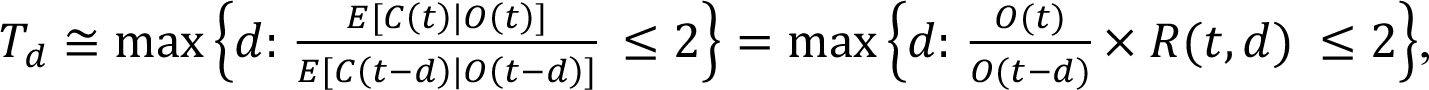

where

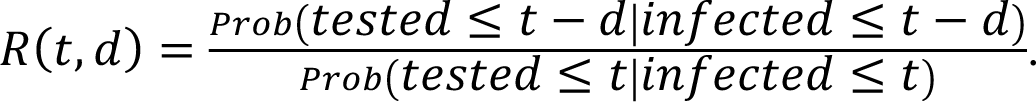

This expression clarifies the limitations in estimating the true doubling time; since we do not know the proportions of infected individuals who are tested at different times, we do not know R(t,d). Nonetheless, it provides us with an understanding of what precisely the estimated doubling time is, and that it is a good estimate of the true doubling time when (t,d) are such that R(t,d) is approximately equal to one. For example, it is reasonable to expect that for t large enough (i.e., enough time into the epidemic) and for d small enough (i.e., short intervals of time), the probability that an infected individual is tested is constant. In particular, if the probability of testing of infected is constant over the d units of time in (t-d,t), this renders the observed doubling time estimate an accurate estimate of the true doubling time. When R(t,d) is larger than one, the crude estimate of the doubling time will be an overestimate, meaning that the course of the epidemic is growing faster than it seems. When it is less than one, the crude estimate is an underestimate, and the epidemic is growing more slowly than it seems.

In some cases, we may be able to estimate R(t,d). Letting D(t) denote the cumulative number of deaths due to the epidemic at time t, we expect the ratio, D(t+*l*)/C(t) to be constant over time, regardless of testing strategy and of the evolution of the epidemic, for some lag time, *l* (3,4). That is, the lethality of the infection should not change over time, regardless of the state of the epidemic. This assumption would be violated if there were more deaths at the height of the epidemic due to limited resources. If this is not the case,

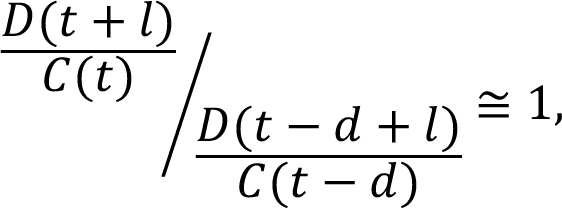

and thus

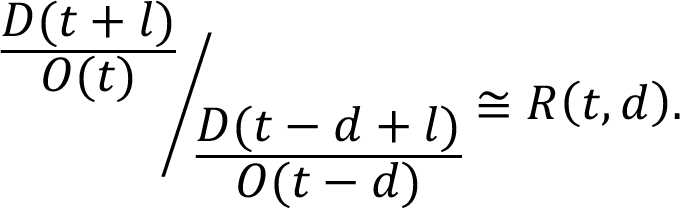

The lag time, *l*, acknowledges the delay from testing to death (4,5); in the case of COVID-19, it has been suggested that *l* should be 18 days, the estimated mean time from symptoms to death (6,7,8). It is important that *all* deaths due to the epidemic be captured in the count D(t), and not just those confirmed by testing. These data have not been reported across the US until recently (9), though they became available for New York City in mid-March (https://github.com/nychealth/coronavirus-data), and so we are able to estimate R(t,d) for New York City. A limitation of this path to the estimation of R(t,d) is that it can be applied only to parameters from *l* days prior to the current time, due to the necessity of the lag.

When counts of probable deaths are not available, precluding estimation of R(t,d), we can conduct sensitivity analyses to determine the robustness of this assumption. In particular, we could assume that in the recent past, the probability of testing of infected might have decreased on day t-s, but otherwise remained constant. This is a conservative assumption since its effect is to increase R(t,u) above 1 for u≤(s-1) and for it to remain equal to 1 for u>s.

An alternative, exploratory analysis is a visualization of the cumulative incidence curve, anchored to a recent date, t^*^, such as 9 days prior to the current date. Using the observed counts for this amounts to a plot of O(t)/O(t^*^) versus t, for t>t^*^. Using the same reasoning as above,

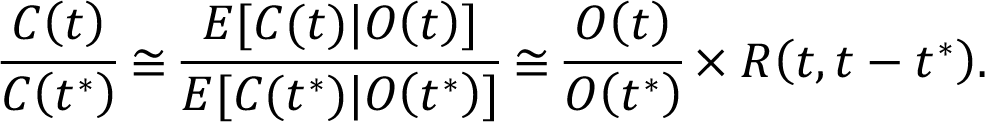

If *R*(*t*,*t* − *t*^*^) ≅ 1, the observed relative cumulative incidence curve provides a good estimate of the true relative epidemic curve. If this assumption is not plausible within (t^*^,t), then alternative values for *R*(*t*,*t* − *t*^*^) can be used in sensitivity analysis.

We have estimated the doubling times and cumulative incidence curves nonparametrically. Parametric alternatives are possible, ranging from log-linear Poisson regression with offset terms to account for testing probability estimates, to fully specified epidemic models. The nonparametric approach is simplest and appropriate when it is desirable that the data fully guide the estimation. An imposed linear assumption has immediate consequences on estimation, and may or may not be accurate.

## Results

Using current data on the number of positive tests in the United States and territories (covidtracking.com) we have estimated the current (April 7, 2020) doubling times for the 12 states with the most cases (Fig. 1). The red “x” indicates the estimate under the assumption that the probability of testing infecteds has not changed over the past few days (2–8 for the states depicted), i.e., R(t,d)≅1. We linearly interpolated the discrete-time estimates. The curves depict a range of sensitivity analyses around worst-case scenarios of over-estimation of the doubling time. Specifically, we calculated doubling times under the assumption that the probability of testing of infecteds was constant in the past, subsequently decreased on a single day in the past, and sustained that decrease through the current time, i.e., R(t,d)>1 for some value of d. The decreases depicted are 10% (green), 20% (blue) and 25% (purple). Of note, the current estimates of doubling time that do not account for potential changes in testing of infecteds might overestimate the true doubling time by as many as three days.

**Fig. 1.**
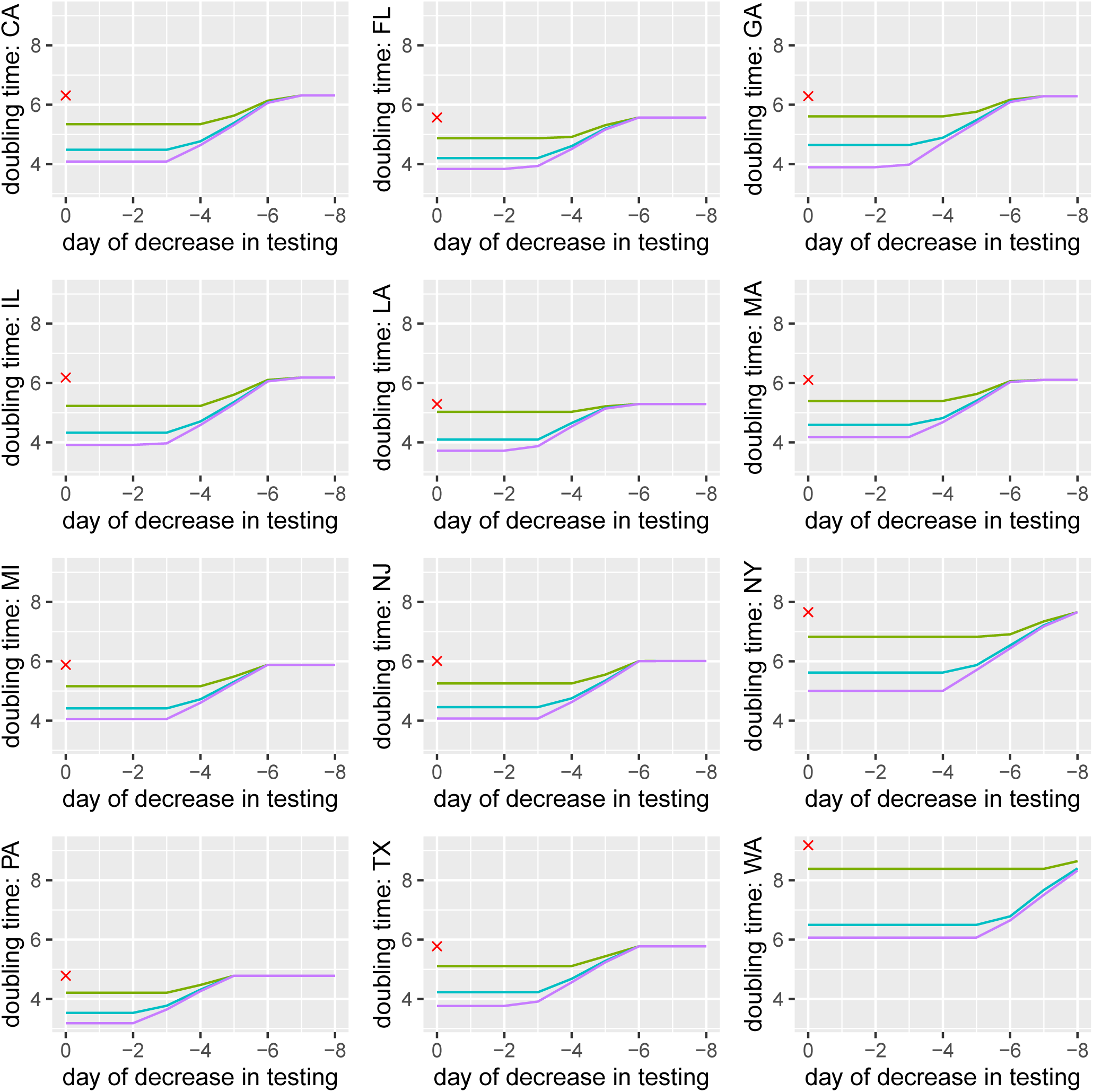
Doubling times with sensitivity to testing for the 12 states with the most cases as of April 7, 2020 (“day 0”). The red “x” is the doubling time estimate on April 7, 2020. The curves depict the current doubling time under the scenario that the probability of testing of infecteds was constant in the past, subsequently decreased on a single day in the past, and sustained that decrease through the current time. The x-axis represents the day on which the probability of testing decreased. The decreases depicted are 10% (green), 20% (blue) and 25% (purple).

In Fig. 2, we display the interpolated doubling times as a function of day to show how they have increased over time. Again, these estimates display sensitivity to potential changes in testing of infected individuals. The red curves assume stable probability of testing over the 20 days considered, the green curves assume a decrease of 10% in probability of testing infected individuals on each current day versus past days, the blue curves assume a decrease of 20% and the purple curves assume a decrease of 25%. This provides another view, over time, of the potential for overly optimistic estimates of doubling time if testing is not considered.

**Fig. 2.**
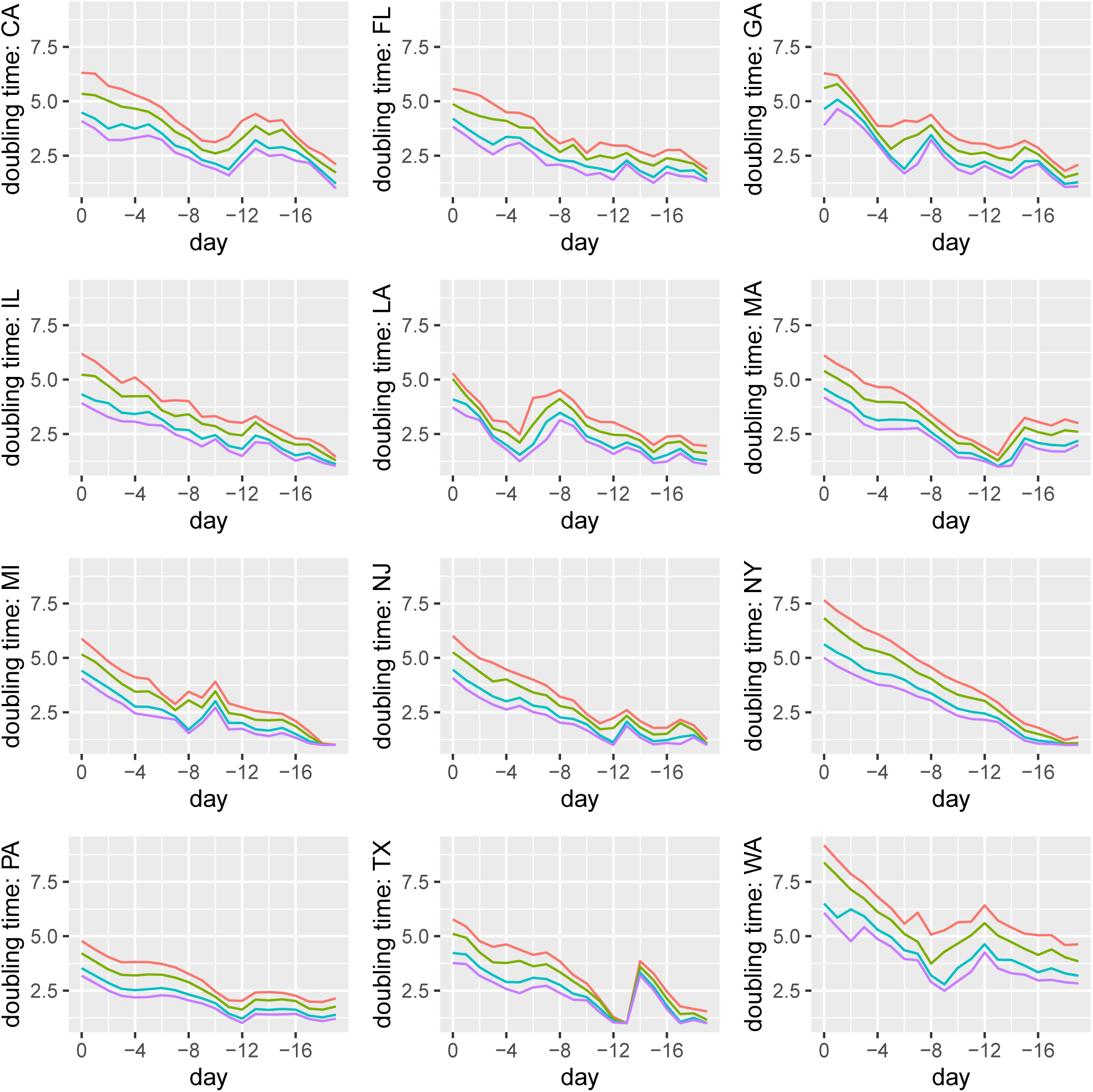
Doubling time on each day from April 7, 2020 back to March 17, 2020. The red curves assume stable probability of testing over the 20 days considered, the green curves assume a decrease of 10% in probability of testing infected individuals on each current day versus past days, the blue curves assume a decrease of 20% and the purple curves assume a decrease of 25%.

In Fig. 3, we display the cumulative incidence curves, standardized by the number of cases identified 9 days prior on March 29, 2020. The red curve assumes that testing of infected individuals has not changed in this timeframe. The colored curves represent increases in the probability of testing of infecteds on each current day. These curves demonstrate that the standardized epidemic curve is subject to underestimation, as a function of testing probabilities.

**Fig. 3.**
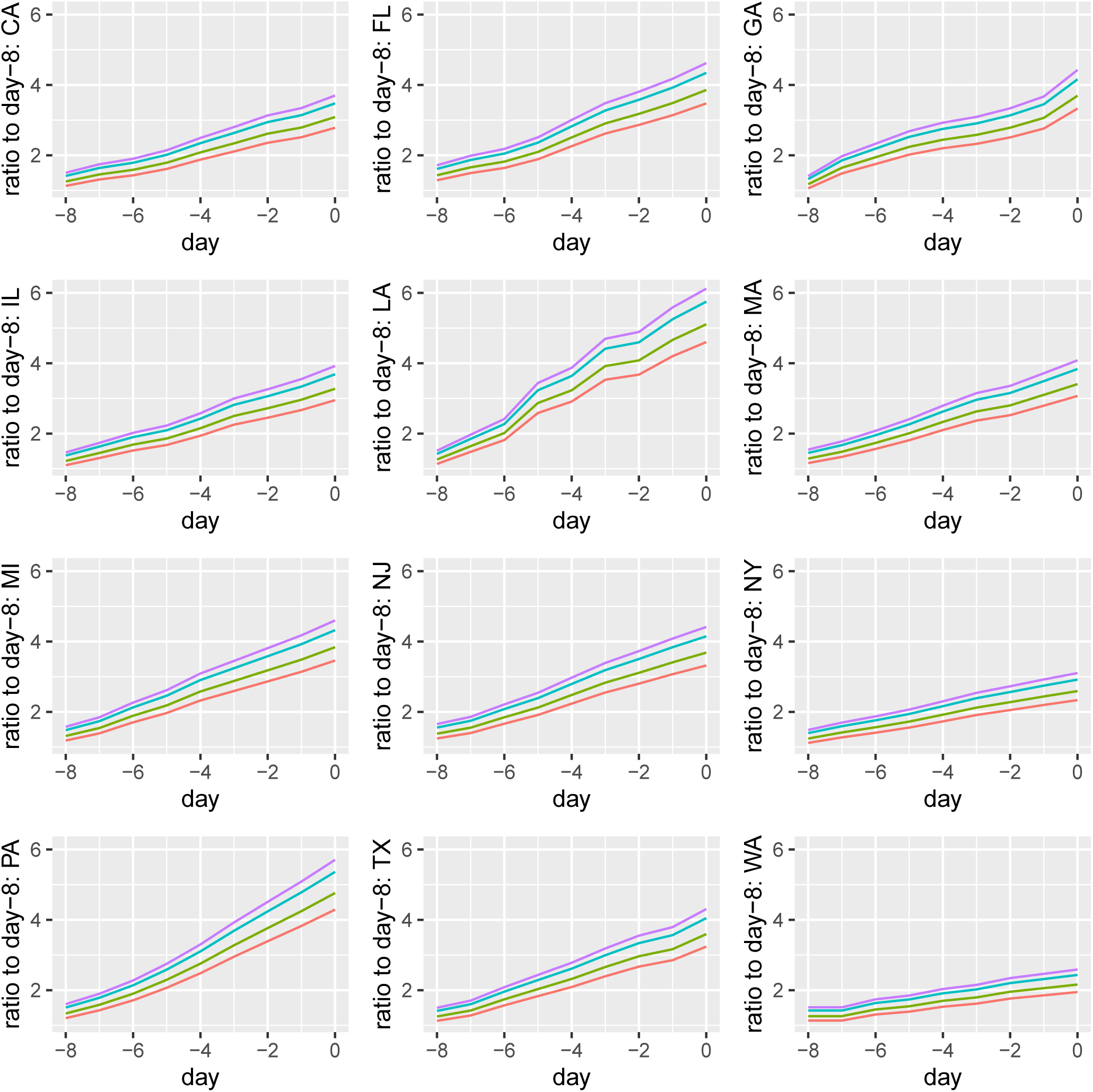
Cumulative incidence curves, standardized by the number of cases identified 9 days ago on March 29, 2020. The red curve assumes that testing of infected individuals has not changed in this timeframe. The green curves assume a decrease of 10% in probability of testing infected individuals on each current day versus March 29, 2020, the blue curves assume a decrease of 20% and the purple curves assume a decrease of 25%.

This series of sensitivity analyses demonstrates that a decrease in testing of infected individuals can lead to an overly optimistic view of the epidemic. In fact, this did not appear to happen in New York City in recent weeks. Fig. 4 depicts R(t,d) as a function of d for New York City, with each curve representing a value of t, ranging from March 25, 2020 through April 7, 2020 and for a lag time, *l*, of 18 days. Over this time period, R(t,d), d=1,…,11, appears to be less than one, which indicates that estimates of doubling time might be smaller, i.e., more pessimistic, than they should be. This indicates also that testing of infected individuals may have increased over time in New York City. A caveat regarding these conclusions is that these estimates are imperfect due to relatively small numbers and likely incompleteness in the numbers of probable deaths. Furthermore, given the necessity of the lag of 18 days, this approach cannot be used for real-time estimation of epidemic parameters.

**Fig. 4.**
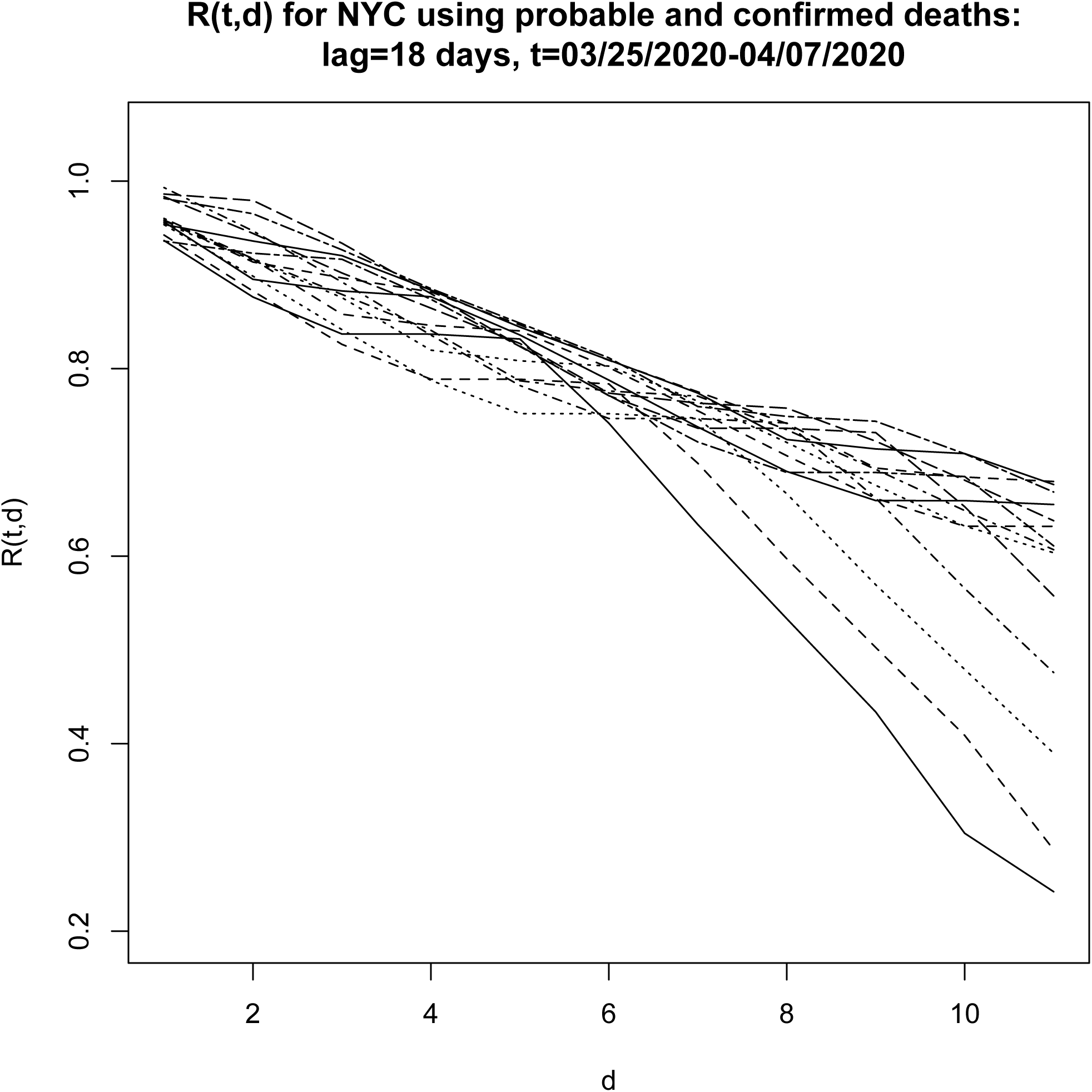
Estimates of R(t,d) for New York City, for t ranging from March 25, 2020 through April 7, 2020 and for lag time, *l*, equal to 18 days.

## Conclusion

In summary, we have illustrated precisely how the nature of testing among infected individuals affects the estimation of important epidemic parameters, which are used to evaluate the utility of public health interventions in communities. In fact, in New York City, the testing of infecteds appears to have increased slightly and so crude estimates may underestimate the slowing of the epidemic. This may not be the case in other states or regions. For this reason, it is important to undertake sensitivity analyses around possible values of R(t,d) to understand its effects on estimates of decreasing doubling times and epidemic curves, and to appropriately evaluate the effects of public health interventions.

## Data Availability

Data are available at covidtracking.com and https://github.com/nychealth/coronavirus-data

https://covidtracking.com/data

## Acknowledgments: Funding

This work was supported by the US National Institutes of Health (R01NS094610) and the US National Science Foundation (DMS-2013789);

## Author contributions

RAB conceived of the study following prior analysis of doubling time by YF. RAB analyzed the data. RAB and YF wrote and approved the manuscript;

## Competing interests

The authors declare no competing interests.

## Data and materials availability

Data were downloaded from covidtracking.com on April 7, 2020 and from https://github.com/nychealth/coronavirus-data on April 26, 2020 and are available in the supplementary materials. The R code used to generate the figures is available in the supplementary materials.

## Supplementary Materials

Materials and Methods

## Notes

### Competing Interest Statement

The authors have declared no competing interest.

